# Effects of temperature on COVID-19 transmission

**DOI:** 10.1101/2020.03.29.20044461

**Authors:** Shrikant Pawar, Aditya Stanam, Mamata Chaudhari, Durga Rayudu

**Affiliations:** Department of Genetics, School of Medicine, Yale University, New Haven, Connecticut, 30303, USA; Department of Toxicology, University of Iowa, Iowa City, Iowa 52242-5000, USA; Department of Biological Sciences, Western Kentucky University, Bowling Green, Kentucky, 42101-1080, USA; Department of Surgery, NTR University, AP, 520008, India

**Keywords:** COVID19, Virus, Death, Recovery, Climate

## Abstract

Coronavirus disease 2019 (COVID-19) is an infectious disease caused by severe acute respiratory syndrome coronavirus 2 (SARS-CoV-2), it was first identified in 2019 in Wuhan, China and has resulted in the 2019–20 coronavirus pandemic. As of March 1, 2020, 79,968 patients in China and 7169 outside of China had tested positive for COVID-19 and a mortality rate of 3.6% has been observed amongst Chinese patients. Its primary mode of transmission is via respiratory droplets from coughs and sneezes. The virus can remain viable for up to three days on plastic and stainless steel or in aerosols for upto 3 hours and is relatively more stable than the known human coronaviruses. It is stable in faeces at room temperature for at least 1-2 days and can be stable in infected patients for up to 4 days. Heat at 56°C kills the SARS coronavirus at around 10000 units per 15 minutes. Thus, temperature is an important factor in survival of COVID-19 virus and this article focuses on understanding the relationship between temperature and COVID-19 transmission from the data available between January-March 2020.

## Introduction

Coronavirus disease 2019 (COVID-19) is an infectious disease caused by severe acute respiratory syndrome coronavirus-2 (SARS-CoV-2) [1]. Ever since, the disease was first identified in 2019 in Wuhan, China, it has spread worldwide quickly. The World Health Organization (WHO) declared the 2019-2020 coronavirus outbreak a pandemic and a Public Health Emergency of International Concern (PHEIC). As of March 20, 2020, more than 240,000 cases have been reported, and more 10,000 people have lost their lives [2]. When the SARS outbreak happened in 2002-2003, SARS-CoV spread to approximately 30 countries only [3]. However, the current outbreak of COVID-19 by SARS-CoV-2 has spread to 176 countries or territories which suggests the enhanced ability of viruses to be aerosolized [2]. Environmental factors such as atmospheric temperature modulates the survival and spread of virus aerosols. It was shown that survival of influenza viral aerosols is reduced at higher temperatures [4]. In this study, we aimed to ascertain the relation between atmospheric temperatures and the COVID-19 disease recovery rates across the globe.

## Methods

### i) Data Collection

The data used in this study comes from Chinese medical community website DXY reporting COVID-19 cases at the province level in China [5]. The data for countries and regions outside mainland China is collected from an interactive web-based dashboard to track COVID-19 in real time [6]. DXY updates case counts every 15 minutes in all provinces in China while other countries’ case counts are manually updated in a web-based dashboard from centres for disease control and prevention (CDC) of Taiwan, Europe, the World Health Organization (WHO), the government of Canada, and the Australian government [6].

### ii) Regression for understanding correlations

The location of confirmed, death and recovered cases was obtained from the coordinate points through NGS coordinate conversion and transformation tool (NCAT) [7]. The respective temperature information for each location was obtained from Raspisaniye Pogodi (RP) temperature channel, Petersburg, Russia having license for the activity in hydrometeorology and adjacent fields. The website provides six day temperature forecasts and information on the actual temperature, observed at ground stations where the forecasts are completely updated twice a day (05:30 and 17:30 UTC) [8]. Temperature reports were downloaded for each location for the day range of 01/22/2020-03/16/2020 which were correlated with the number of confirmed positive cases, deaths and recoveries. The *ggpairs()* function from the GGally package was used to create a plot matrix to see how the variables relate to each other [9]. The coefficient errors provide a variation of the estimated coefficients from the actual average while the t-value measures how many standard deviations the estimated coefficient is from zero. Residuals describe how well does the model fit the data and a bell curve distribution is an ideal for any linear fit [10]. Residuals, coefficients, degrees of freedom, R-squared and F-statistic values are provided in Table 1.

**Table 1:**
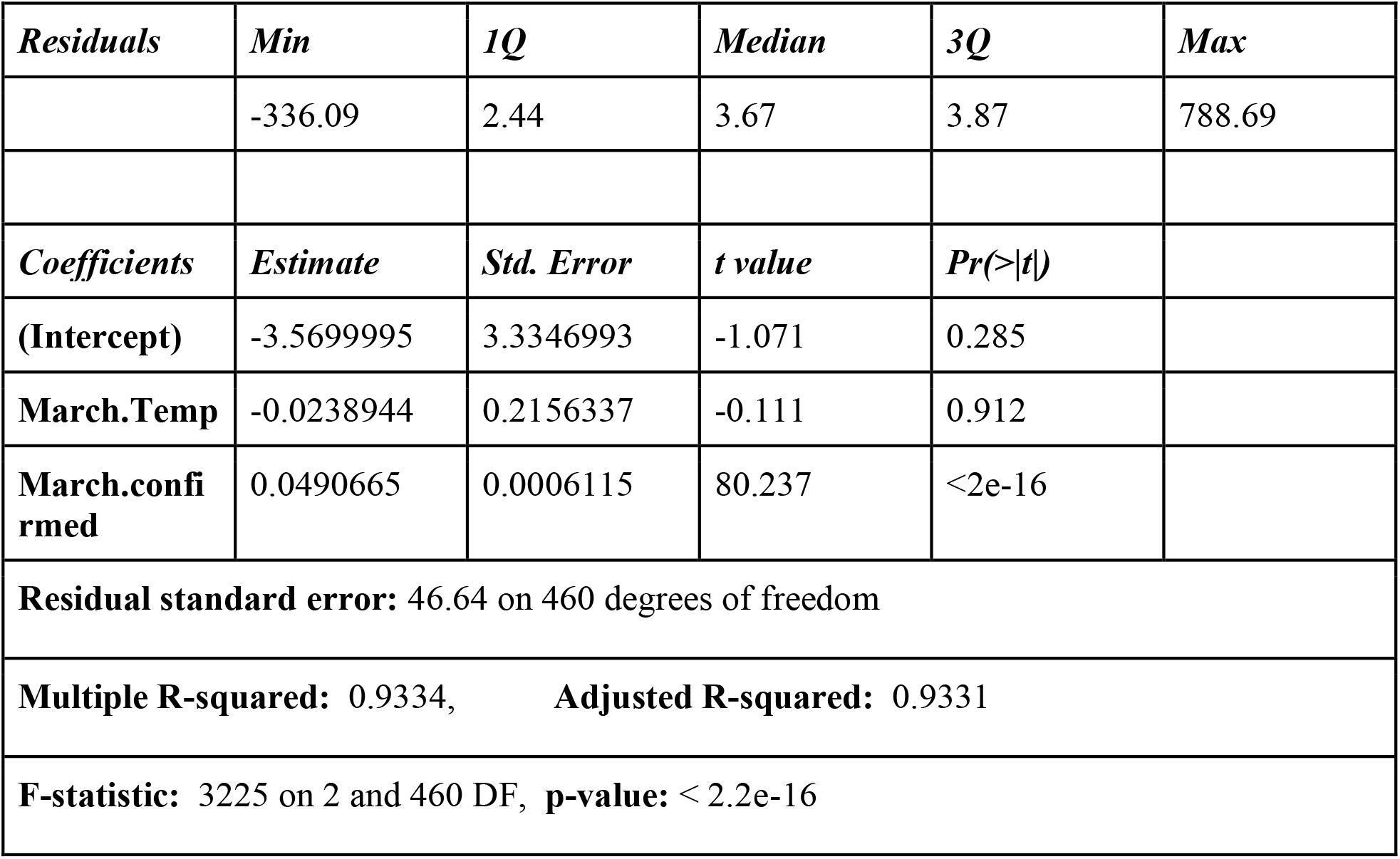
Residuals, coefficients, degrees of freedom, R-squared and F-statistic for multiple regression models.

## Results

### i. Countries including China, Iran, Italy, Germany and France showed maximum impact with COVID-19 transmission and morbidity

The data retrieved from 01/22/2020-03/16/2020 showed 81033 confirmed positive cases of COVID-19 infection in China, 27980 in Italy, 7272 in Germany, 6650 in France and 14991 cases in Iran (Figure 1 & supplementary file 1). In the same period of time, China reported 3217 deaths, while France, Iran, Italy and Spain reported 148, 853, 2158, and 342 deaths respectively (Figure 2 & supplementary file 1). China, Italy and Iran were highly impacted by COVID-19 mortality. A speedy recovery was also seen in China with 67910 cases, while Iran and Italy with 4590 and 2749 cases recovered by 03/16/2020 (Figure 3 & supplementary file 1). Hubei province in China was hit worse with 3099 deaths followed by Henan and Heilongjiang provinces with 22 and 13 deaths respectively (Figure 4, Left), while the state of California was severely affected in the United States of America (US) with 7 deaths reported (Figure 4, Right).

**Figure 1:**
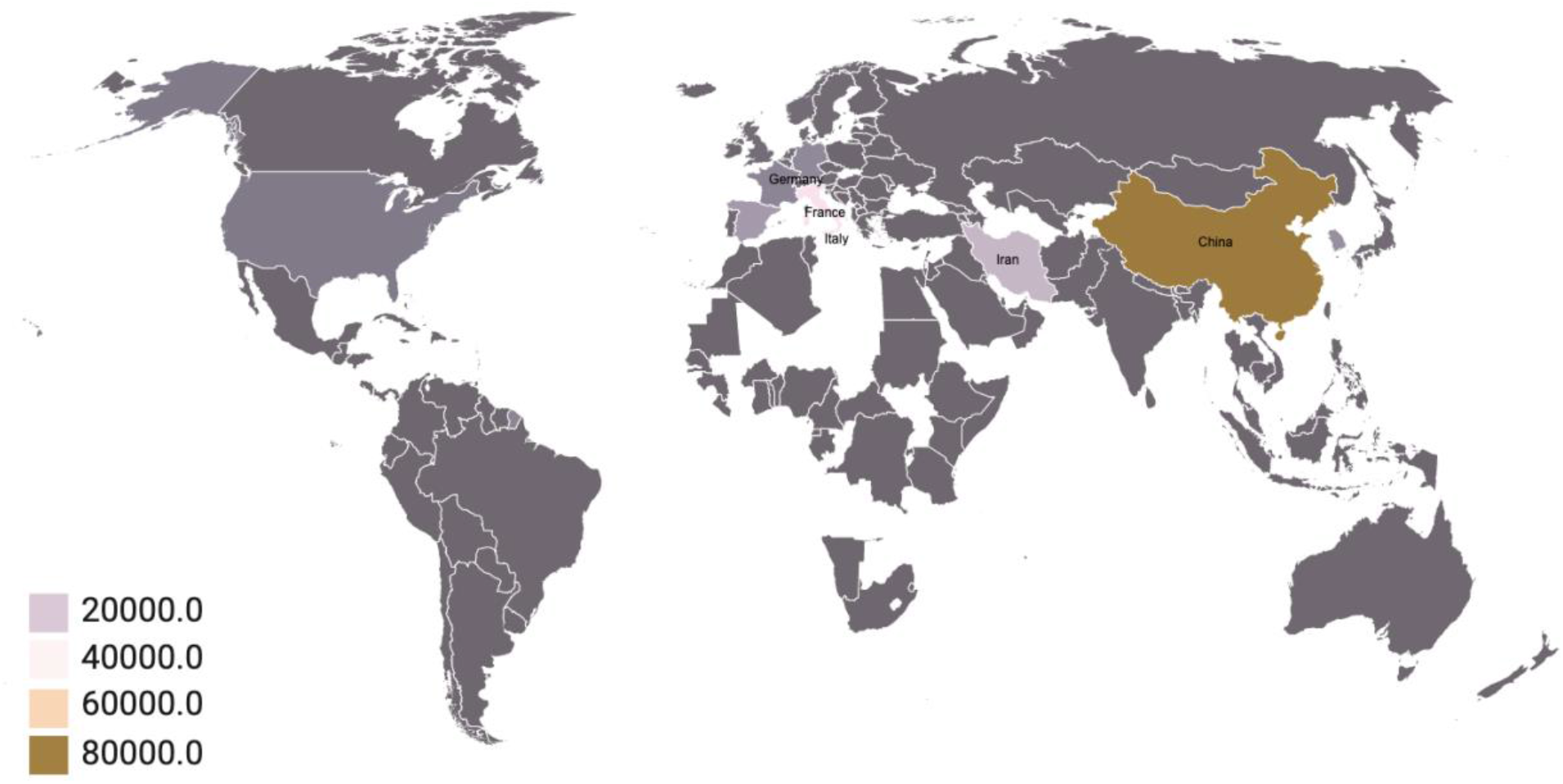
World map of cases tested confirmed positive with COVID-19 infection. There were 81033 confirmed positive cases of COVID-19 infection in China, 27980 in Italy, 7272 in Germany, 6650 in France and 14991 cases in Iran.

**Figure 2:**
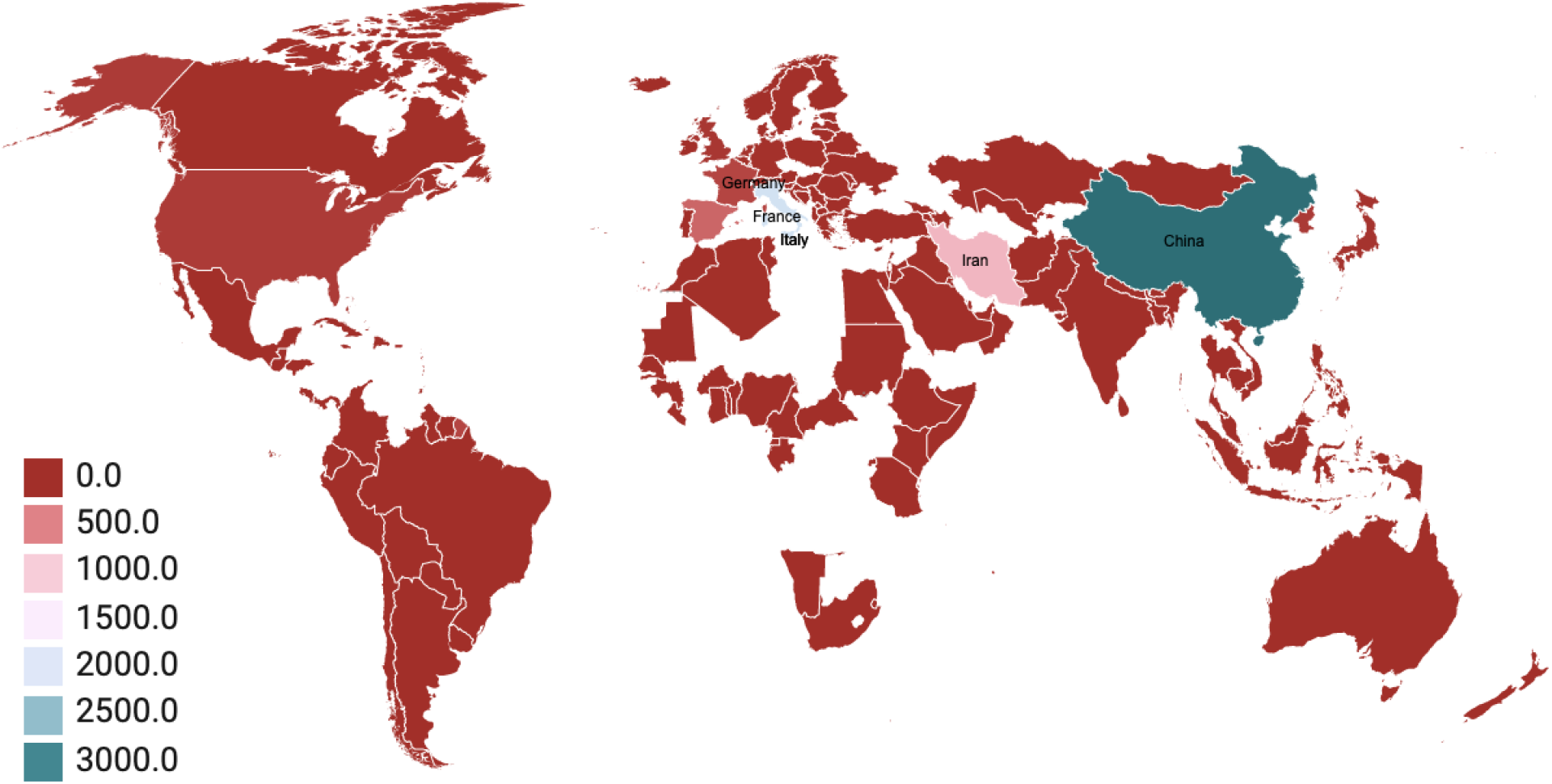
World map of cases confirmed deaths with COVID-19 infection. China reported 3217 deaths, while France, Iran, Italy and Spain reported 148, 853, 2158, and 342 deaths respectively.

**Figure 3:**
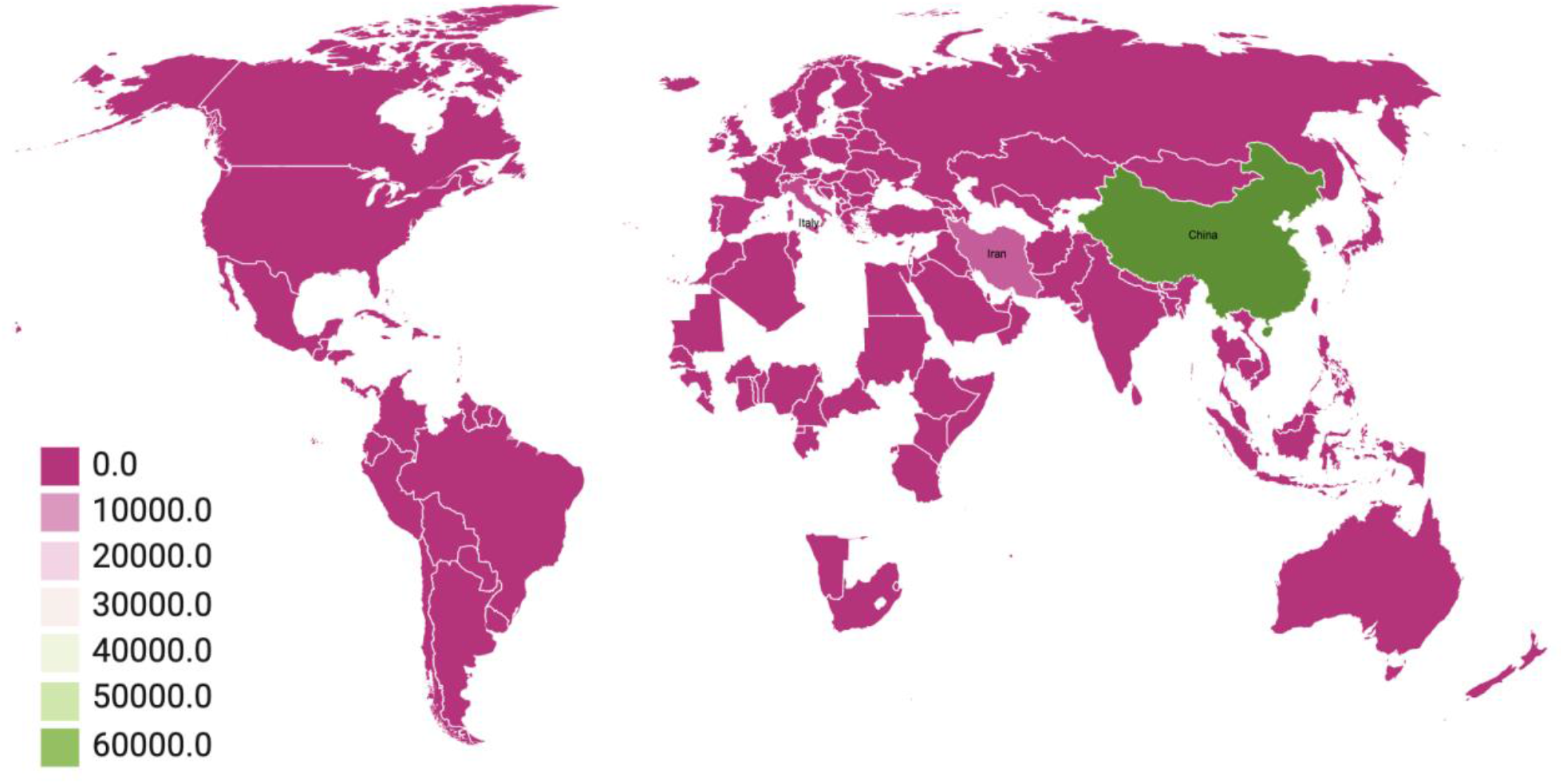
World map of cases confirmed recovered with COVID-19 infection. Around 67910 cases were recovered in China, while Iran and Italy recorded 4590 and 2749 cases recovered by 03/16/2020.

**Figure 4:**
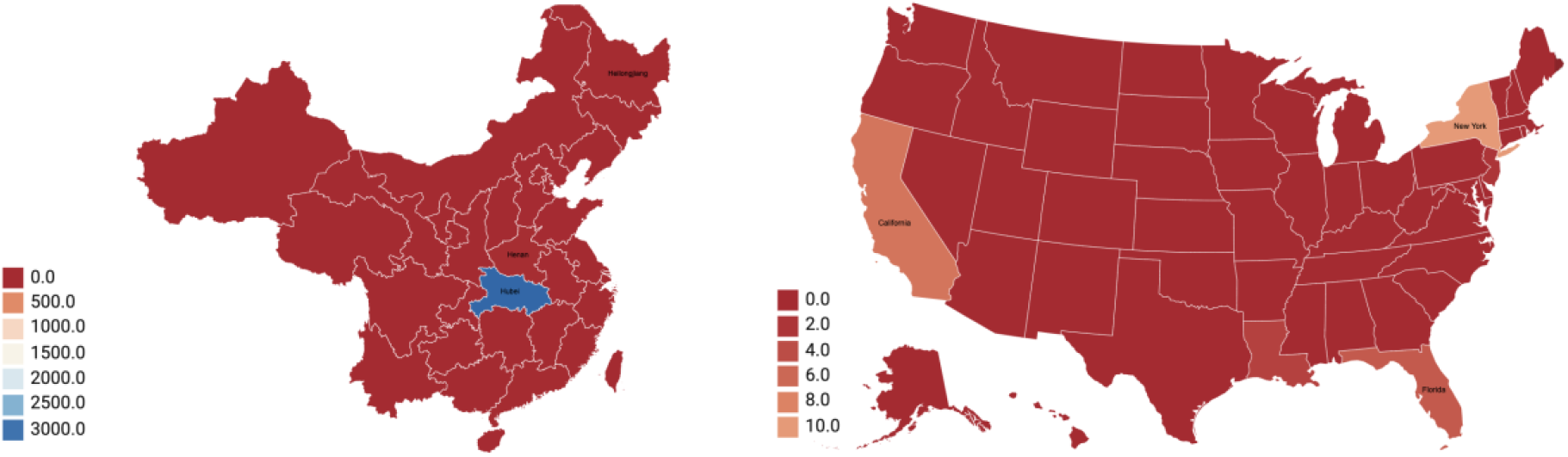
Map of People’s Republic of China (PRC) (Left) & United States of America (Right) for cases confirmed deaths with COVID-19 infection. Hubei province in China was hit worse with 3099 deaths followed by Henan and Heilongjiang provinces with 22 and 13 deaths respectively (Figure 4, Left), while California state was severely affected in the United States of America with 7 deaths (Figure 4, Right).

### ii. Changes in temperature shows no significant correlation with cases transmitted, deaths or recovered

Linear regression describes the relationship between a response variable of interest and one or more predictor variables by separating the signal from the noise [11]. Correlation analysis between temperatures, cases confirmed positive, dead and recovered was performed separately for overall countries in the month of January, February and March of 2020. For cases confirmed positive, dead and recovered, a total number of cases was used as a variable value while an average of temperature (degree celsius (°C)) for each of the months was used as a variable temperature value. Comparing overall cases for temperature, no significant correlation between temperatures and cases confirmed positive, dead or recovered was observed (Figure 5A), however, a strong correlation (∼0.9) was observed between cases confirmed and deaths for each of the three months. To further investigate effects of temperature, country (Australia) with highest average temperature (January∼18°C, February∼17°C, March∼16°C) was compared separately with the country (Canada) with lowest average temperature (January∼-3°C, February∼-5°C, March∼-2°C) for the month of March 2020 (Figure 5B: Top Left and Top Right). Again, no significant correlation between temperatures and cases confirmed positive, dead or recovered was observed, however, an interesting correlation pattern was seen where a drop in correlation (∼0.5) was found between cases confirmed and recovered in Australia and a drop in correlation (∼0.3) was found between cases confirmed and deaths in Canada. China being the earliest and highly affected country with COVID-19 transmission, separate correlation plots were generated between temperatures and cases confirmed positive, dead or recovered for the months January-March 2020 (Figure 5B: Bottom Left, Bottom Right; Figure 5C: Left). While no significant correlation between temperatures and cases confirmed positive, dead or recovered was observed, a strong correlation between cases confirmed and recovered (∼0.9), and cases confirmed and deaths (∼1) was recorded. Finally, a correlation matrix (Figure 5C: Right) for the month of March 2020 for the US was generated and again no significant correlations were observed between any of the selected variables.

**Figure 5:**
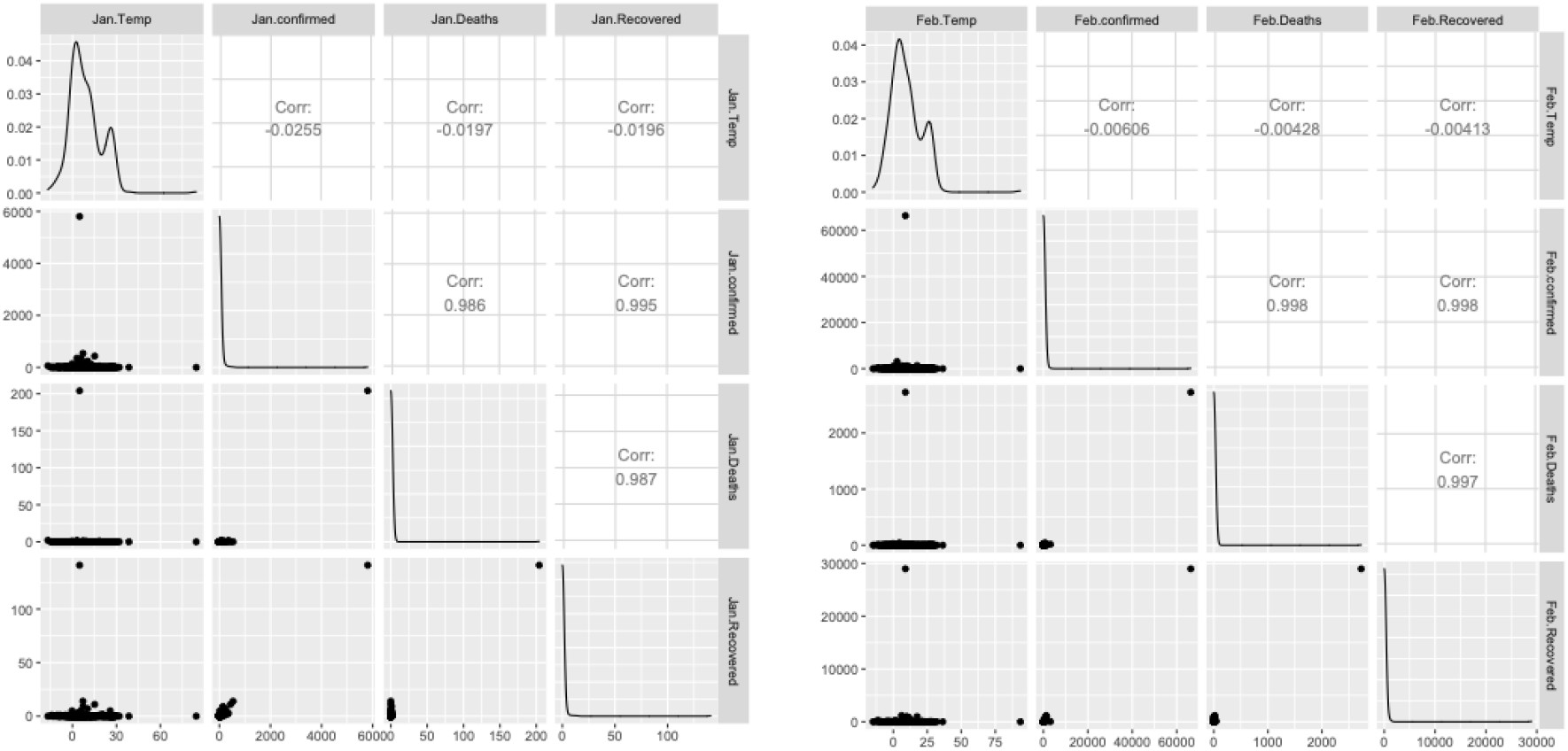
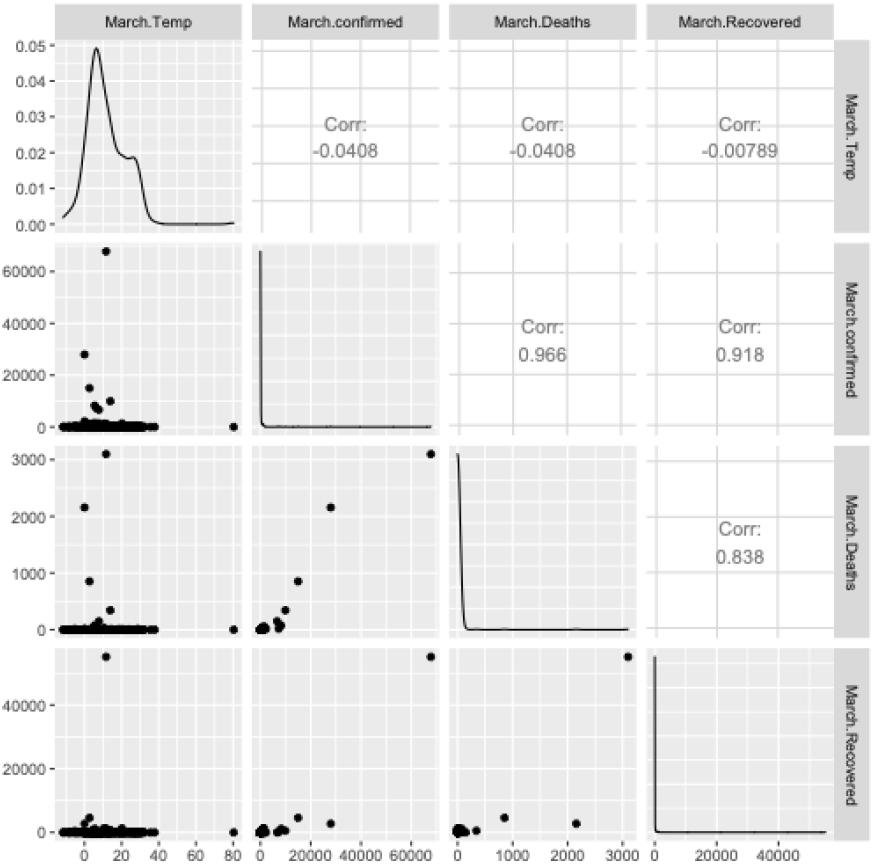
Scatter plots for each variable combination with density plots for each variable and the strength of correlations between variables. Figure 5A: Top Left: Overall correlation matrix for month of January 2020. Top Right: Overall correlation matrix for month of February 2020. Bottom Center: Overall correlation matrix for month of March 2020.

**Figure 5B:**
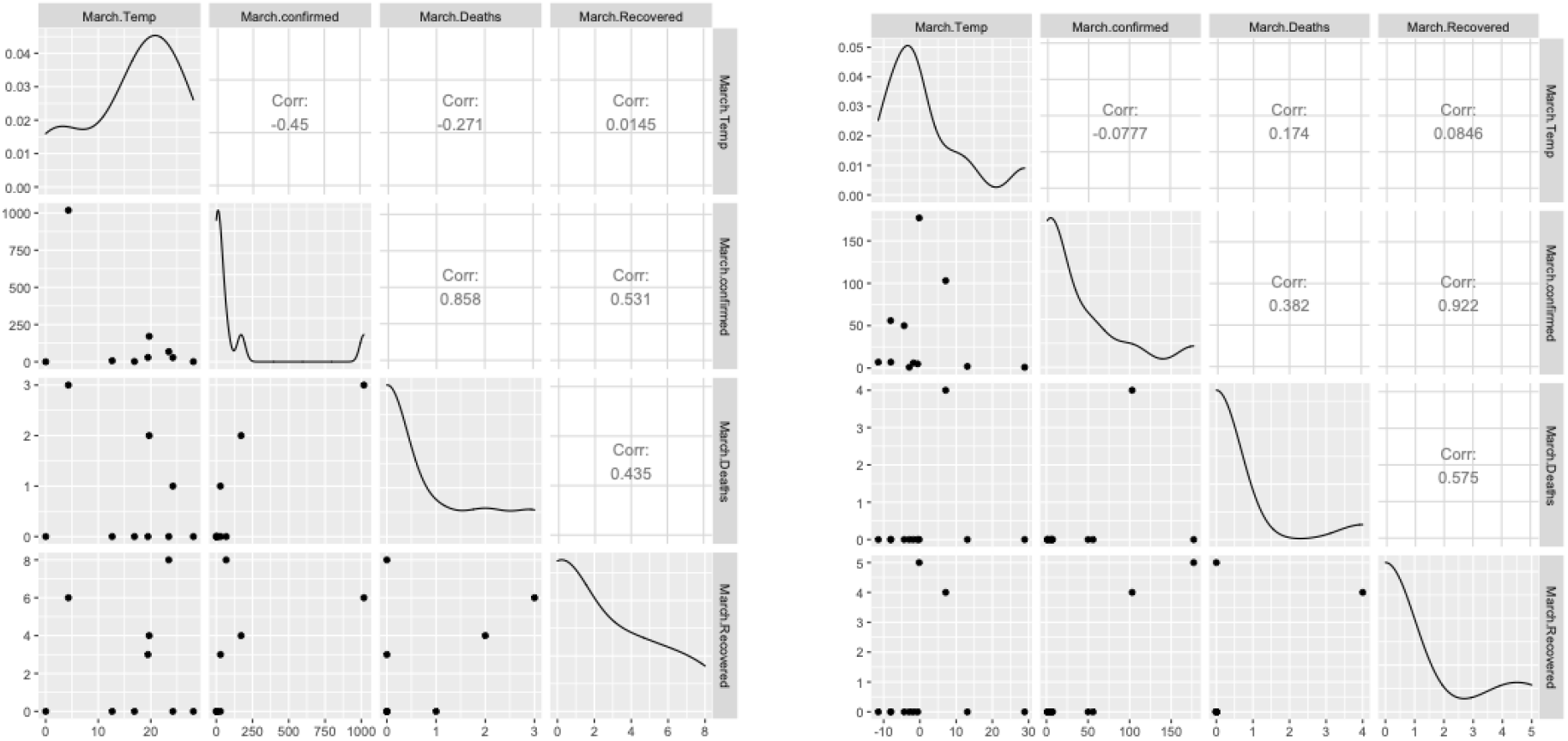
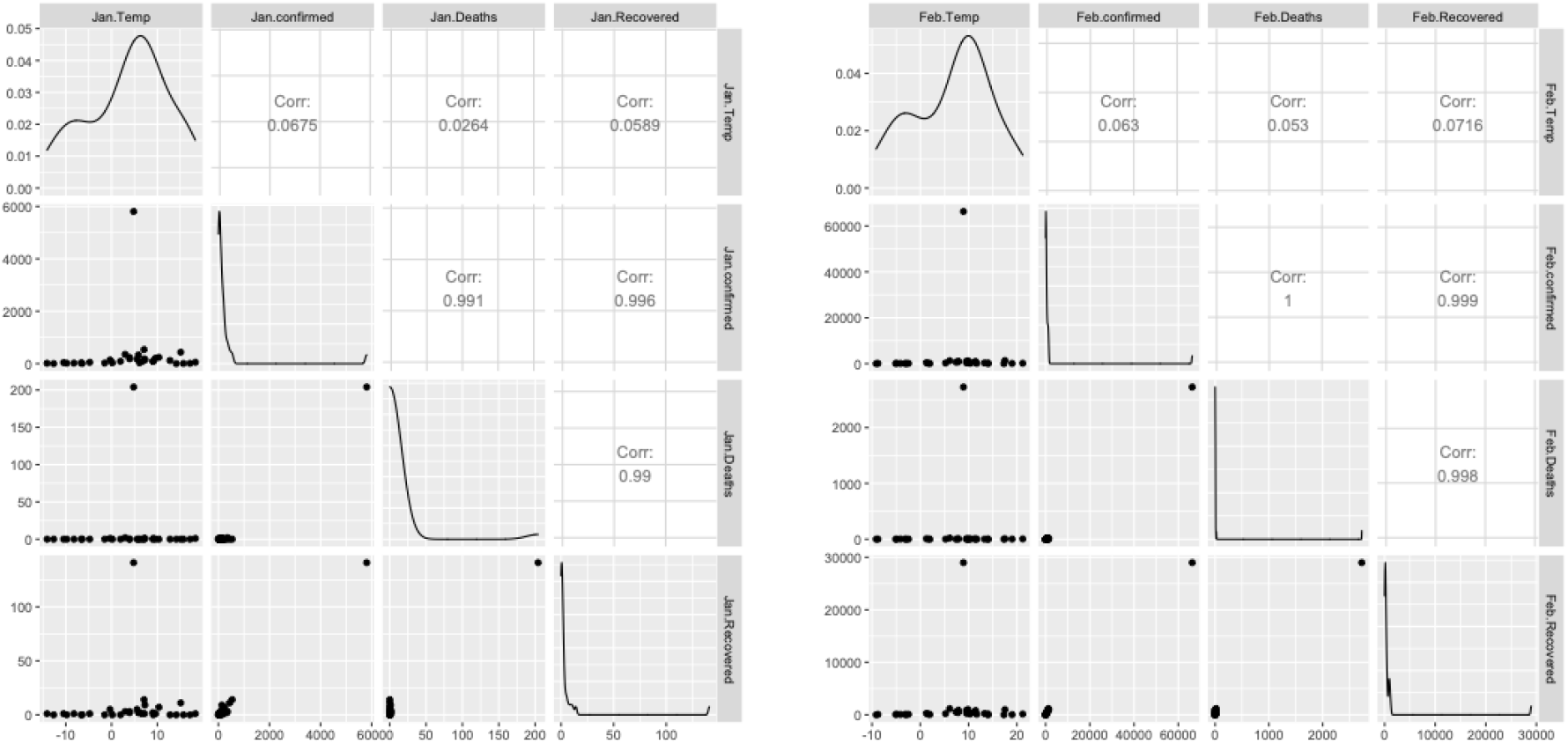
Top Left: Correlation matrix for month of March 2020 for Australia. Top Right: Correlation matrix for month of March 2020 for Canada. Bottom Left: Correlation matrix for month of January 2020 for China. Bottom Right: Correlation matrix for month of February 2020 for China.

**Figure 5C:**
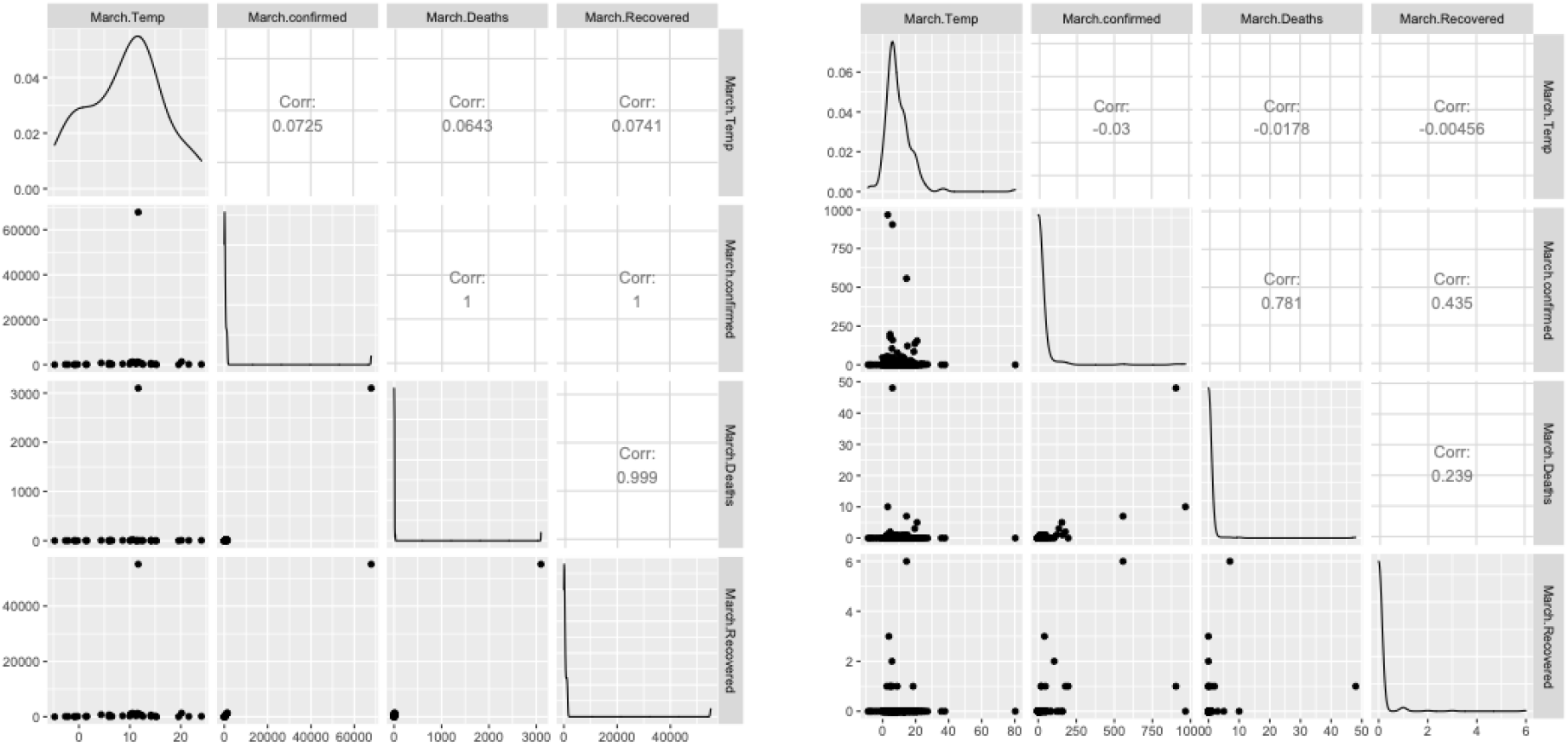
Left: Correlation matrix for month of March 2020 for China. Right: Correlation matrix for month of March 2020 for the US.

### iii. Regression model predicts rise in number of deaths in China, US Australia and Canada

A multiple regression analysis was performed to create a model for predicting deaths from cases confirmed and an average temperature of certain locations. A strong correlation between cases confirmed with deaths and cases confirmed with recovered was observed, so a robust regression model was expected. Although, we found insignificant correlations between temperatures and cases confirmed positive, dead or recovered, considering it as one of the variables in multiple regression model won’t make a significant difference. A significant P value and F score was obtained (Table 1 and Figure 6: Left) stating robustness of the generated model. For the month of March 2020, Australia reported a total of 377, Canada reported 415, China reported 81033 and the US reported a total 4632 confirmed positive cases. The average temperatures of Australia, Canada, China, and the US in March was ∼16, −0.2, 8.4 and 9.2°C respectively. Considering a 1°C rise on each of these temperatures for April 2020 and combining them with their respective confirmed positive case numbers for March 2020, predictions for the number of deaths were made from the regression model (Figure 6: Right).

**Figure 6:**
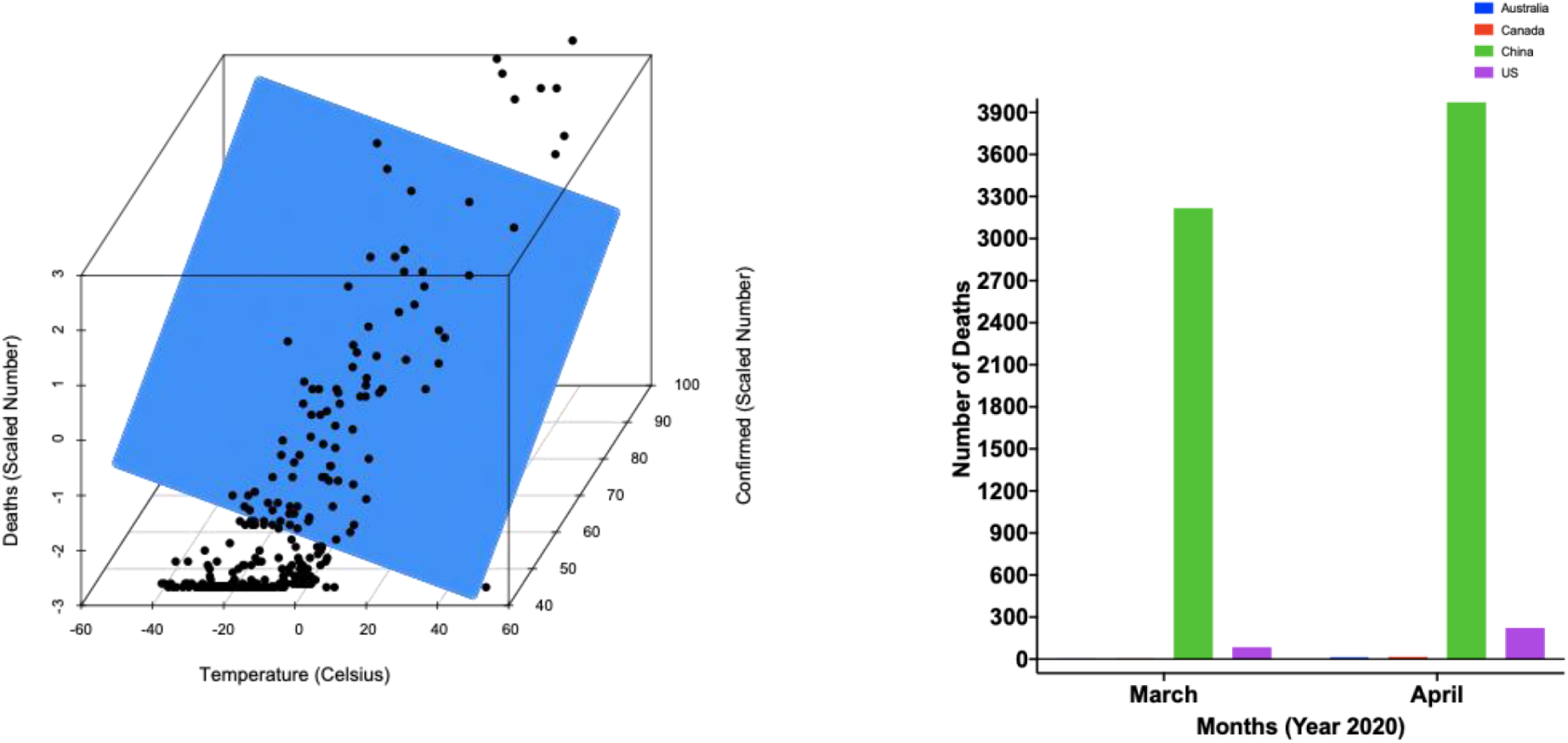
(Left) 3d scatterplot from the predictor grid and the predicted volumes (Blue grid), original data (Black dots). (Right) Bar plot for predicted number of deaths for different countries.

## Discussion

Data analyzed from 01/22/2020-03/16/2020 for COVID-19 confirmed positive, death and recovered cases identify an increasing number of cases being recovered (∼83%) especially in China, while Iran and Italy showed a slow recovery with 30% and 37% recovery rates respectively. China reported the highest percent deaths (∼3.96%) in the world. Although a strong correlation between cases confirmed with recovered, and cases confirmed with deaths was observed, changes in temperature showed no significant correlation with cases transmitted, deaths or recovered for the period 01/22/2020-03/16/2020. However, an interesting finding was observed wherein a drop in correlation was found between cases confirmed with recovered in Australia (average temperature ∼16°C) and cases confirmed with deaths in Canada (average temperature ∼-2°C). Finally, a multiple regression model was generated using predicted April 2020 temperatures with confirmed positive cases for March 2020 for Australia, Canada, China, and the US to predict the number of deaths in April 2020. The generated model predicted ∼14, 16, 3972 and 223 more deaths for countries Australia, Canada, China, and the US respectively. This data and model signifies urgency of confining COVID-19’s pandemic spread with the predicted rise in number of deaths.

## Data Availability

The data used in this study is not publicly available, but can be obtained by emailing the corresponding author.

## Author contributions

SP and AS conceived the concepts, planned and designed the article. SP and AS primarily wrote and edited the manuscript. DR and MC supported the data collection and analysis.

## Supplementary Files

**File 1: Overall number of cases positive, dead and recovered from 01/22/2020-03/16/2020**.

**File 2: Number of cases positive, dead and recovered with latitude, longitude and average temperatures from 01/22/2020-03/16/2020 divided within 3 months**.

